# Identification of predictive patient characteristics for assessing the probability of COVID-19 in-hospital mortality

**DOI:** 10.1101/2023.07.16.23292738

**Authors:** Bartek Rajwa, Md Mobasshir Arshed Naved, Mohammad Adibuzzaman, Ananth Y. Grama, Babar A. Khan, M. Murat Dundar, Jean-Christophe Rochet

## Abstract

As the world emerges from the COVID-19 pandemic, there is an urgent need to understand patient factors that may be used to predict the occurrence of severe cases and patient mortality. Approximately 20% of SARS-CoV-2 infections lead to acute respiratory distress syndrome caused by the harmful actions of inflammatory mediators. Patients with severe COVID-19 are often afflicted with neurologic symptoms, and individuals with pre-existing neurodegenerative disease have an increased risk of severe COVID-19. Although collectively, these observations point to a bidirectional relationship between severe COVID-19 and neurologic disorders, little is known about the underlying mechanisms. Here, we analyzed the electronic health records of 471 patients with severe COVID-19 to identify clinical characteristics most predictive of mortality. Feature discovery was conducted by training a regularized logistic regression classifier that serves as a machine-learning model with an embedded feature selection capability. SHAP analysis using the trained classifier revealed that a small ensemble of readily observable clinical features, including characteristics associated with cognitive impairment, could predict in-hospital mortality with an accuracy greater than 0.85 (expressed as the area under the ROC curve of the classifier). These findings have important implications for the prioritization of clinical measures used to identify patients with COVID-19 (and, potentially, other forms of acute respiratory distress syndrome) having an elevated risk of death.

## 1 Introduction

### 1.1 COVID-19 and neurologic symptoms

Patients with severe COVID-19 warranting hospital admission present with a variety of symptoms that are carefully evaluated by admitting physicians. These patient evaluations have led to the identification of specific comorbidities linked to severe forms of COVID-19 or fatal outcomes [1–3], including neurodegenerative disorders and dementia [4–6]. Although SARS-CoV-2 infection can cause neurologic symptoms by directly affecting the central nervous system (CNS), this phenomenon has only been shown in a very small subset of patients [7]. Instead, neurologic symptoms are more likely to occur due to indirect effects involving the strong innate immune response and cytokine storm caused by SARS-CoV-2 infection.

A cytokine storm is a hyperinflammatory response to an infection caused by a sudden spike in levels of pro-inflammatory cytokines and chemokines, including IL-1, IL-2, IL-4, IL-6, IL-7, IL-8, IL9, IL-10, IL-18, granulocyte stimulating factor (G-CSF), IP-10, monocyte chemoattractant protein (MCP)-1, MCP-3, macrophage inflammatory protein 1 (MIP-1A), cutaneous T-cell attracting chemokine (CTACK), IFN-γ, and TNF-α). In turn, this phenomenon can result in overwhelming systemic inflammation, acute respiratory distress syndrome (ARDS), and multi-organ failure [8]. Evidence suggests that a cytokine storm can trigger various neurologic symptoms, ranging from headaches, dizziness, and disorientation to convulsions or seizures [8].

### 1.2 Approach to prediction and feature selection

In this report, we present the results of a study aimed at establishing a link between COVID-19 symptoms observed upon patient admission (within the first 24 h of hospitalization) and the risk of patient death. We were particularly interested in the predictive value of easily observable neuropsychiatric symptoms such as disorientation, cognitive impairment, and delirium. Our strategy involved a data-driven discovery of predictors. Instead of postulating *a priori* a feasible set of clinical features likely to be associated with mortality and then testing the resulting hypotheses using a standard generalized linear model (GLM) approach, we retained all possible clinical features for the analysis. We used the disease outcome to train a supervised classifier with feature ranking and selection capability. When an explainable classifier achieved high accuracy, we queried it to determine which feature combination was responsible for its strong performance. Finally, we hypothesized that the predictive combinations of clinical features were causally linked to mortality or that common latent factors influenced both the mortality as well as the detected clinical characteristics.

We utilized two independent approaches to quantify the predictive power of the observations collected at the time of patient admission, or within the first 24 h of hospitalization: elastic-net regularized logistic regression (LR-ENET) and XGBoost classification. The use of both methods was followed by an analysis of the relative contributions of the selected features (“explanatory variables”) to the prediction via the SHapley Additive exPlanations (SHAP) approach. Both methods led to the identification of several important predictive features, including neurologic symptoms.

### 1.3 Related research

Data-driven investigation of COVID-19 mortality employing machine learning tools (such as XGBoost) and feature explanation methods (SHAP values) has been demonstrated before for the processing of clinical laboratory results [9–11], for predicting death outcomes [12–14], demonstrating links between socioeconomic disparities and COVID-19 spread [15], and showing the impact of COVID-19 on mental health in self-identified Asian Indians in the USA [16]. Several other articles using SHAP were reviewed recently by Bottino et al. [17].

## 2 Methods and datasets

### 2.1 Dataset description

Electronic health record (EHR) data were obtained from 471 patients with severe SARS-CoV-2 infection. The patients were admitted to the intensive care units (ICUs) of IU Health Methodist Hospital and Sidney & Lois Eskenazi Hospital, both in Indianapolis, Indiana. 399 patients were eventually discharged, whereas 72 patients died. The demographic characteristics of the cohort are shown in Table 1. 246 patients self-identified as *Black* or *African American*, and 196 patients identified as *white*. 245 of the patients were females, and 226 were males. There was no statistically significant difference in age between the African-American and white patients. However, patients who identified as Hispanic or Latino were significantly younger than other patients (*p*<0.001).

**Table 1:** Demographic characteristics of the investigated cohort.

| Race | Female |  |  |  |  | Male |  |  |  |  |
| --- | --- | --- | --- | --- | --- | --- | --- | --- | --- | --- |
|  | Alive | Alive Perc. | Died | Died Perc. | Total | Alive | Alive Perc. | Died | Died Perc. | Total |
| Asian | 3 | 100.0% | 0 | 0.0% | 3 | 5 | 83.3% | 1 | 16.7% | 6 |
| Black or Afr. Amer. | 123 | 87.9% | 17 | 12.1% | 140 | 85 | 80.2% | 21 | 19.8% | 106 |
| Refused to identify | 1 | 33.3% | 2 | 66.7% | 3 | 2 | 100.0% | 0 | 0.0% | 2 |
| Unknown | 10 | 100.0% | 0 | 0.0% | 10 | 5 | 100.0% | 0 | 0.0% | 5 |
| White | 77 | 86.5% | 12 | 13.5% | 89 | 88 | 82.2% | 19 | 17.8% | 107 |
| Total | 214 | 87.3% | 31 | 12.70% | 245 | 185 | 81.9% | 41 | 18.1% | 226 |

**Table 2:** Mean ages, Braden scores, Glasgow coma scores, and probabilities of demonstrating a clear speech pattern for patients in the investigated cohort.

| Sex | Status | Age, mean | Age, IQR | Braden score, mean | Coma score, mean | $\mathcal{P}(\text{Clear speech})$ | (95% CI) |
| --- | --- | --- | --- | --- | --- | --- | --- |
| Female | alive | 55.08 | 25.61 | 16.92 | 13.80 | 0.84 | (0.79, 0.88) |
|  | died | 80.27 | 19.35 | 13.90 | 12.84 | 0.48 | (0.32, 0.65) |
| Male | alive | 56.70 | 20.18 | 17.11 | 14.14 | 0.82 | (0.76, 0.87) |
|  | died | 75.31 | 12.85 | 14.15 | 13.29 | 0.56 | (0.41, 0.7) |

### 2.2 Preprocessing

The original dataset consisted of data collected at multiple time points during the patients’ treatment. We used only the data from the first time point (i.e., the initial evaluation), consisting of the earliest available diagnostic characteristics. We retained the minimal and maximal values if multiple measurements and/or laboratory results were provided. Because many features describe the patient’s status as very detailed and granular, some binary factors were observed for only a few (one or two) patients. These variance-deficient features were removed to prevent the model from outfitting, even though they might have been informative. Another step of feature engineering (not pursued in this study) is likely needed to construct virtual features that summarize these descriptors.

The presence of multiple correlated features related to delirium is an example of the described problem. We found that the large number of delirium-related descriptors would lead to the emergence of 29 distinct categories. However, because over 300 cases showed no evidence of delirium, an alternative approach would be to combine all of the positive categories into one (e.g., “some evidence of delirium”). Although we did not use this engineered feature in our preliminary model, we examined separately the predictive value of the “delirium” secondary feature. Indeed, as Table 3 demonstrates, patients assigned to the class “no delirium” had a substantially lower probability of death.

**Table 3:** Probability of death given sign of delirium at the hospital admittance and accompanying lower (LCL) and upper (UCL) 95-percentile confidence limits.

| Sex | Status | $\mathcal{P}(\text{Death})$ | LCL | UCL |
| --- | --- | --- | --- | --- |
| Female | No delirium | 0.08 | 0.05 | 0.12 |
|  | Delirium | 0.39 | 0.25 | 0.56 |
| Male | No delirium | 0.13 | 0.09 | 0.19 |
|  | Delirium | 0.46 | 0.30 | 0.62 |

### 2.3 Logistic regression with elastic-net regularization model

We established a set of relevant diagnostic features by implementing an *ante-hoc* explainable, predictable statistical model with embedded feature selection capability. We utilized utilize generalized linear models (GLMs) regularized with a ridge (*ℓ* _2_), LASSO (*ℓ* _1_), or a combination of both penalties (elastic net) [18–20]. This approach allowed us to (1) create a simple model capturing all the significant sources of variability, incorporating all of the diverse clinical descriptors/features, and (2) perform simultaneous feature selection and feature ranking, allowing identification of the major drivers of correct prediction.

The model has been defined as follows:

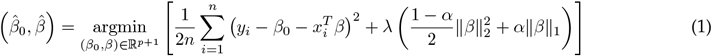

where *λ* is the tuning hyperparameter controlling the overall strength of the LASSO (*ℓ* _1_) and ridge (*ℓ* _2_) penalties, and *α* controls the balance between them. Because the elastic net regularization penalizes the size of the coefficients, sets some irrelevant values to 0, and minimizes the impact of irrelevant features, the feature importance can be expressed straight from the model by the absolute values of the non-zero coefficients of the covariates.

Unfortunately, minor adjustments in the random initialization or train-test split of the model leads to considerable variances in the selected feature set for the majority of embedded feature selection methods. This problem is known as a selection instability [21, 22]. In general, *ℓ* _1_ regularization in GLMs is known to be unstable [23]. However, investigating ensemble feature selection, in which the set of optimal features is produced from a collection of multiply independently trained models, helps resolve this issue. There are multiple approaches to increase the feature selection stability by performing various implementations of the ensemble approach, most notably the RENT model, which combines information about the frequency of feature occurrence and feature weights [21]. We followed a simple, yet effective, approach of training 10 independent models, each of which was initiated with a different random seed. To account for the data imbalance, we used the ROSE (Random OverSampling Examples) algorithm [24]. The simulated instances for training were generated de novo for each independent model.

### 2.4 XGBoost model

The extreme gradient boosting decision tree (XGBoost), developed by Chen and Guestrin, is a highly effective, portable, and scalable machine learning system for tree boosting that is optimized under the Gradient Boosting framework [25]. It combines a series of low-accuracy weak classifiers using the gradient descent architecture to produce a strong classifier with higher classification performance. For a dataset *D* = (*x*_*i*_, *y*_*i*_) : *{i* = 1…*n, x∈* ℝ, *y∈* ℝ*}* with n samples and m features, the predicted value 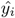 of the XGBoost model can be represented as:

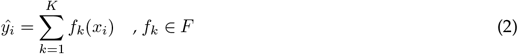

where *f*_*k*_ represents a CART tree and the score given by the *k*-th tree to *i*-th data sample is denoted by *f*_*k*_(*x*_*i*_). The set of K such functions is learned by minimizing the following objective function:

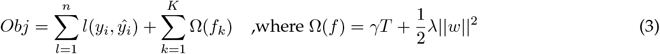

Here, *l* is a convex training loss function that measures the difference between prediction 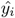and target *y*_*i*_; Ω is a model complexity function term that penalizes the complexity of the XGBoost model, where *γ* and *λ* are degrees of regularization. *T* and *w* refer to the number of leaves and the scores on each leaf of the tree, respectively. The XGBoost model can be trained in an additive manner. Given 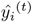 as the prediction of the *i*^th^ instance at *t*^th^ iteration, function *f*_*t*_ needs to be added to minimize the objective function:

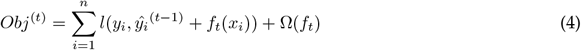

By applying Taylor expansion this function is simplified as:

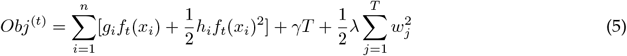

where *g*_*i*_ and *h*_*i*_ are the first and second derivatives obtained on the loss function, respectively. By calling the stated tree creation model repeatedly, a large number of regression tree structures are acquired. The objective function, *Obj*, is then used to choose the optimal tree structure and insert it into the existing model to create the optimal XGBoost result.

### 2.5 SHapley Additive exPlanations (SHAP) values

The machine learning field has adopted the cooperative game theory concept known as SHAP (or Shapley) values [26–29]. The SHAP values can determine the importance of a feature and its directionality influence by comparing what a model predicts with and without that feature for each observation in the training data and calculating the marginal contribution [27].

Briefly, the exact Shapley values are computed based on the following procedure. Let’s consider an M-player cooperative game in which the objective is to maximize the payoff, and let 𝒮*⊆M* = *{*1, …, *M}* be a subset of |𝒮| players. Further, let’s assume we have a contribution function *v*(𝒮) that maps subsets of players to real numbers, which we refer to as the worth or contribution of coalition 𝒮. The worth of coalition 𝒮 describes the expected total sum of payoffs that the members of can obtain through cooperation [30]. The Shapley value is one method for distributing the total gains to the players, assuming that they are all cooperating. It is a “fair” distribution (i.e., characterized by efficiency, symmetry, null player property, and linearity) and is expressed as:

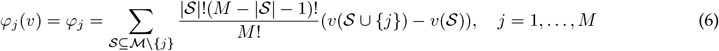

which is the weighted mean over contribution function differences for all subsets of 𝒮players not containing player *j*. Colloquially, the 𝒮values illustrate how important each player is to the overall cooperation and what payoff the player can reasonably expect from participation in the game. SHAP values provide a straightforward way to determine which features contribute to a prediction by considering a model trained on a set of features as a value function on a coalition of players. Importantly, Shapley values may have causal interpretations where the conventional “conditioning by observation” as in Pearl’s do-calculus, can be replaced by “conditioning by intervention” [31, 32].

## 3 Results

### 3.1 Patients characteristics

Patients who died of COVID-19 were significantly older than patients who survived (*p*<0.001) in both the female and male groups. The average Braden score was slightly lower among patients who died (females: *p*=0.035, males: *p*=0.021). There was no observable difference between the mean Glasgow Coma scores of patients who died or survived (females *p*=0.14, males: *p*=0.15). Patients who presented with clear speech and, therefore, presumably had intact cognitive ability were significantly over-represented among those who survived (odds ratio of being admitted with clear speech for surviving patients: 5.65 (*p*<0.001) for females, 3.6 (*p*<0.001) for males). See Table 2 for a summary of the findings.

The composite feature describing the presence of any delirium-related symptoms was an excellent univariate predictor of patient outcomes. Patients exhibiting delirium symptoms had a significantly higher probability of death, a relationship that was evident in both the female and male groups. See Table 3.

To compare the formulated models to a benchmark, we created a simple GLM employing the composite delirium feature, sex, race, Braden score, and age category discretized into three tertiles: *young* [<55.7], *middle* [51.7, 66.8] and *older*[>66.9]. The model is represented as:

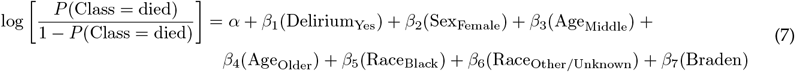

Inspection of the model demonstrates that age is the most important predictive factor, followed by delirium symptoms and the Braden score. There is no significant difference between patients of different races. Table 4 shows the results of the statistical analysis, and 5 presents the average marginal means computed from the model. After fitting, the model has an AUC of 0.87, a sensitivity of 0.94, and a specificity of 0.3. Post-hoc analysis demonstrated that after controlling for race, sex, age group, and the Braden score, the odds of death for patients exhibiting delirium symptoms increased by 5.23 (*p*<0.001).

**Table 4:** Statisitical summary of the model shown in Equation 7.

| Predictor |  |
| --- | --- |
| Class: died/survived | Log odds (SE) |
| Delirium symptoms | 1.655*** (0.339) |
| Female | -0.707** (0.319) |
| Age [51.8, 66.9] | 2.486** (1.058) |
| Age [over 66.9] | 4.440*** (1.028) |
| Black | -0.158 (0.317) |
| Other | 0.108 (0.785) |
| Braden score | -0.043** (0.021) |
| Constant | -4.330*** (1.081) |
| Observations | 471 |
| Log Likelihood | -136.291 |
| AIC | 288.583 |
| R<sup>2</sup> Tjur | 0.282 |

Note: \* $p < 0.1$ ; \*\* $p < 0.05$ ; \*\*\* $p < 0.01$

**Table 5:**
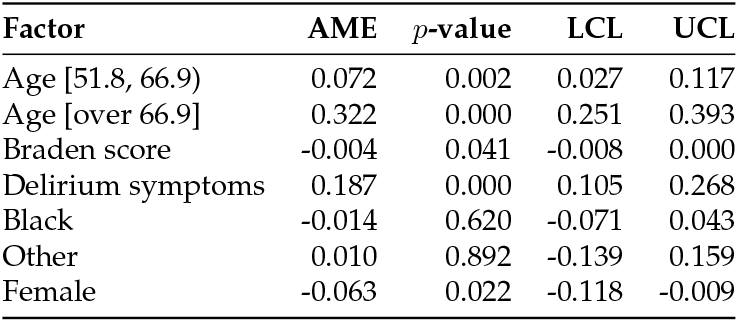
Average marginal means, lower and upper confidence intervals associated with the factors incorporated in the benchmark model introduced in Equation 7.

### 3.2 Logistic regression model results

LR model training was performed using a grid search through the space of parameters *λ* and *α*. An example of a training grid is illustrated in Figure 1.

**Figure 1:**
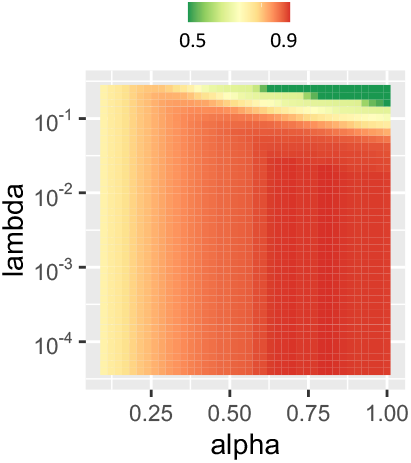
Changes in performance of the elastic-net regularized model in the classification of COVID-19 patients, expressed as AUC, given different values of αand γparameters.

The performance of the trained LR models is demonstrated in Figure 2. The classifier performed well, and was consistently able to reach AUC of approximately 0.9.

**Figure 2:**
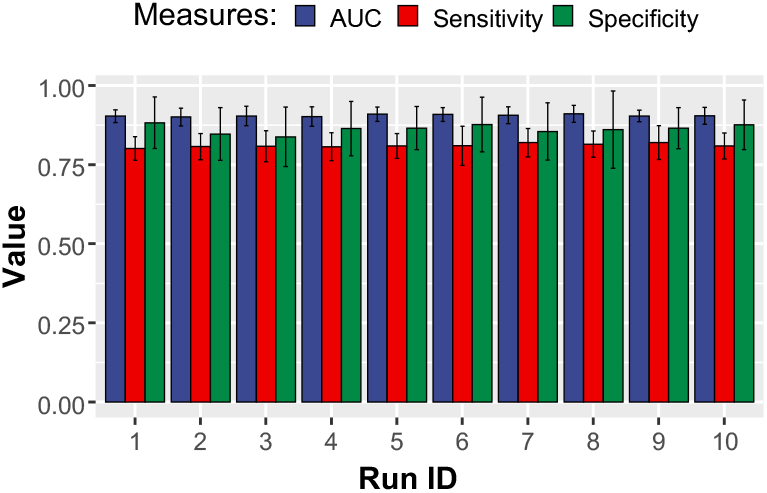
Performance of the elastic-net regularized model in the classification of COVID-19 patients. Ten independent rounds of model training are shown.

Each of the regularized logistic regression models in the ensemble was trained to limit the number of utilized features to approximately 20. For the training and feature selection process, each classifier used the available data augmented with synthetic cases generated using ROSE algorithm [24]. The augmentation step was incorporated to counter any data imbalance. Each of the runs employed a newly synthesized set of points for augmentation, contributing to additional variability that challenged the ensemble classifier. Training cross-validation was performed utilizing the 0.632 bootstrap.

After each run, absolute values of the different LR-ENET models were collected and scaled to [0,1] intervals. These measures were considered reflective of the predictors’ importance. Due to instability, several lower-performing predictors were observed with non-zero coefficients though only in a few runs. On the other hand, consistent predictors emerged in most or all of the runs. All of the scaled predictive importance values were subsequently analyzed, and the top 20 were picked for the final selection.

The results illustrating the identified predictive EHR features are summarized in Figure 3.

**Figure 3:**
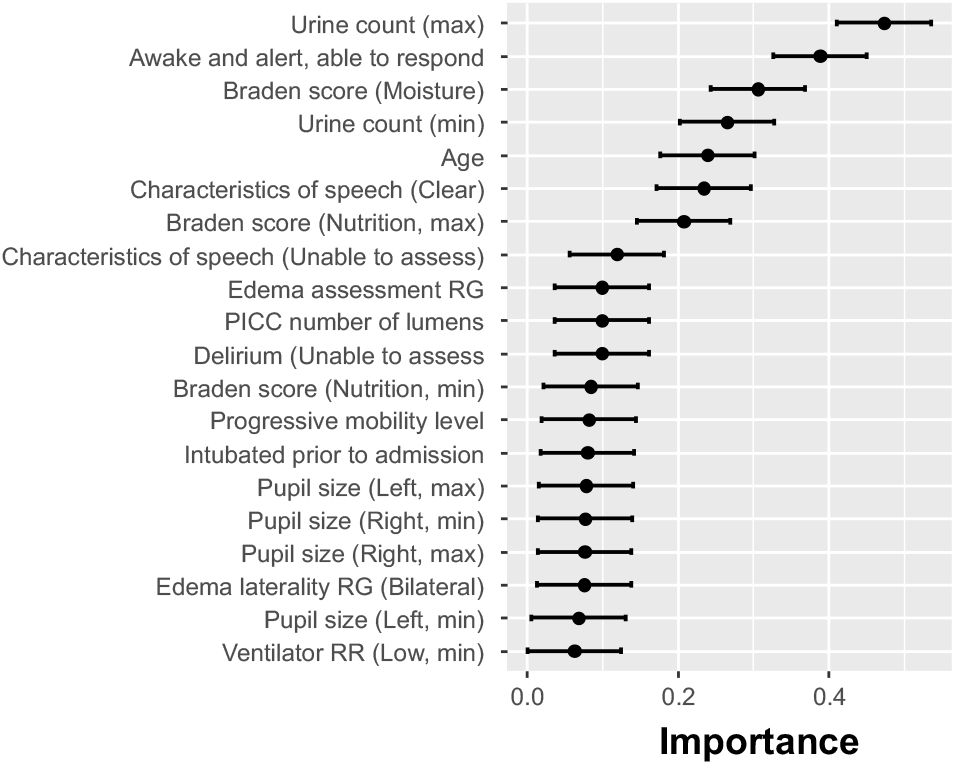
Importance of elastic-net selected features shown as the normalized absolute value of the regression coefficients

As mentioned before, given the instability of sparse classifiers, every run may provide a slightly different list of selected features. Therefore, formally, the SHAP analysis should be performed for every one of the runs. However, to gather some preliminary insight into the explanatory power of the selected features, we ran the SHAP analysis only for one of the ten LR-ENET-trained classifiers. The results are demonstrated in Figure 4.

**Figure 4:**
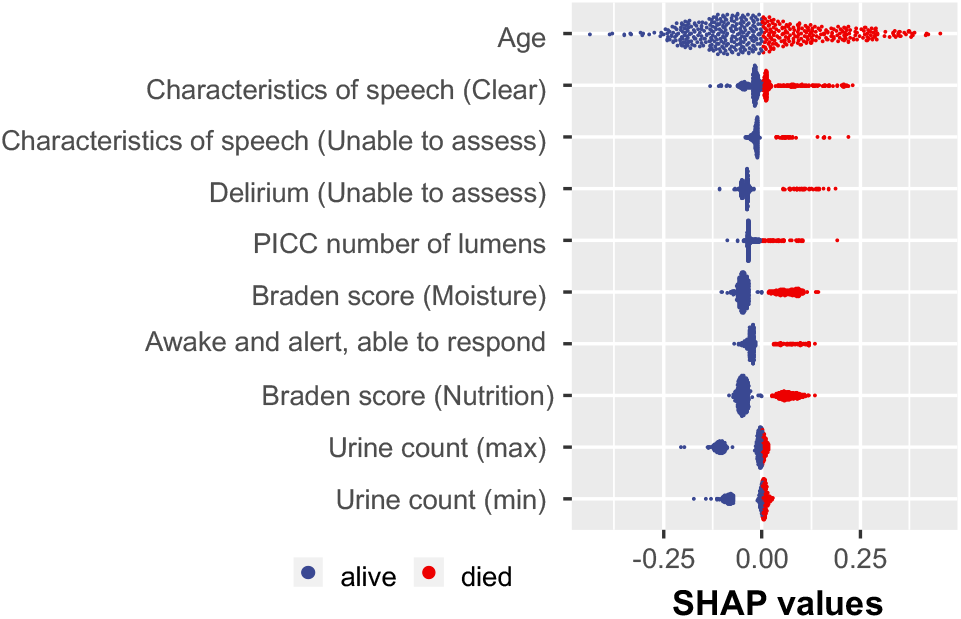
Example of the SHAP value distribution for all of the tested patients and the augmented dataset. The visualization was created based on one of the trained LR-ENET classifiers.

### 3.3 XGBoost model results

XGBoost performed comparably to the LS algorithm, although it is noted that the acquired specificities were often lower. We optimized the XGBoost hyperparameters to maximize the AUC as opposed to explicitly minimizing the number of utilized features. Therefore, the algorithm was allowed to employ as few or as many features as were required to optimize its performance.

The XGBoost model can be used as a feature selection wrapper. In the process of training the features are selected in ignored in the created trees. On the basis of that, the XGBoost method ranks the most significant characteristics according to “Gain,” “Cover,” and “Frequency.” The gain reflects how crucial a characteristic is for making a branch of a decision tree pure. Coverage measures the proportion of observations affected by a feature. A feature’s frequency is the number of times it is used in all created trees (See Figure 5).

**Figure 5:**
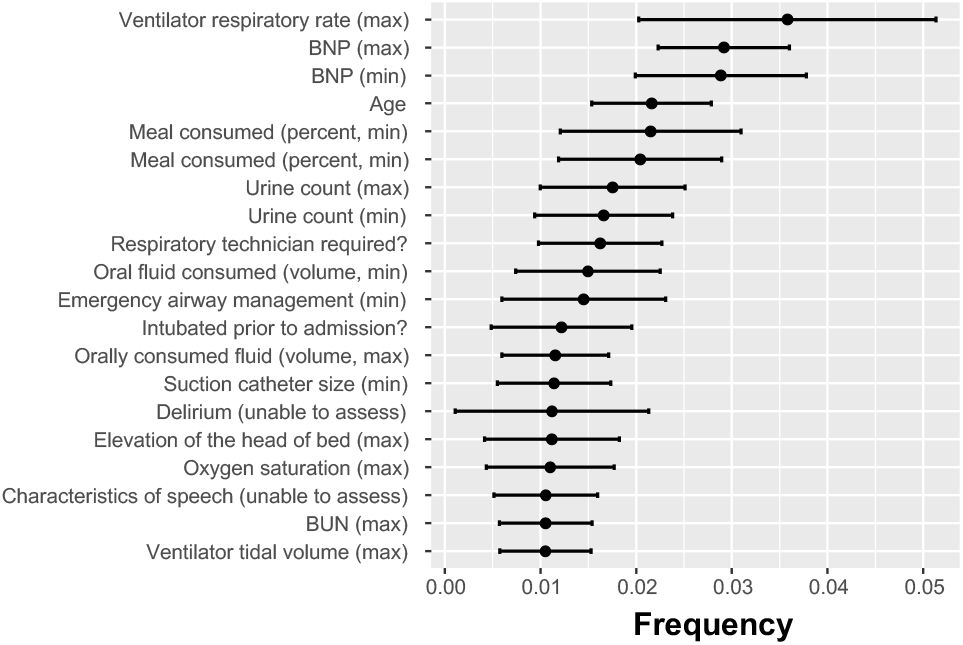
Importance of features employed by XGBoost algorithm expressed as the frequency at which each feature was used in the created classification trees.

The XGBoost algorithm was not directly constrained by the number of used features. The performance of the XGBoost classifier is shown in Figure 6. The XGBoost-discovered features were exposed to the SHAP analysis, the results of which are demonstrated in Figure 7.

**Figure 6:**
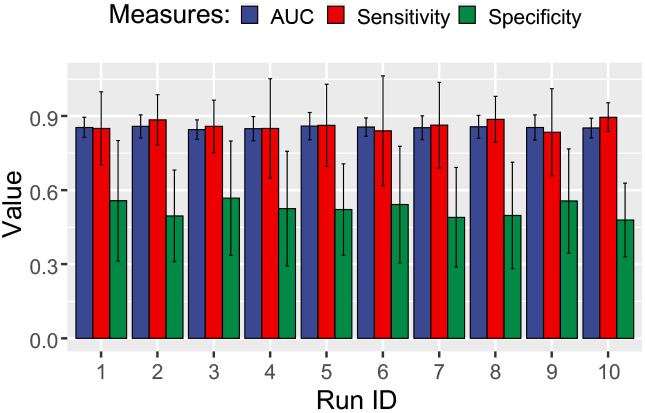
Performance of the XGBoost model in repeated independent cross-validation rounds classifying the COVID-19 patients.

**Figure 7:**
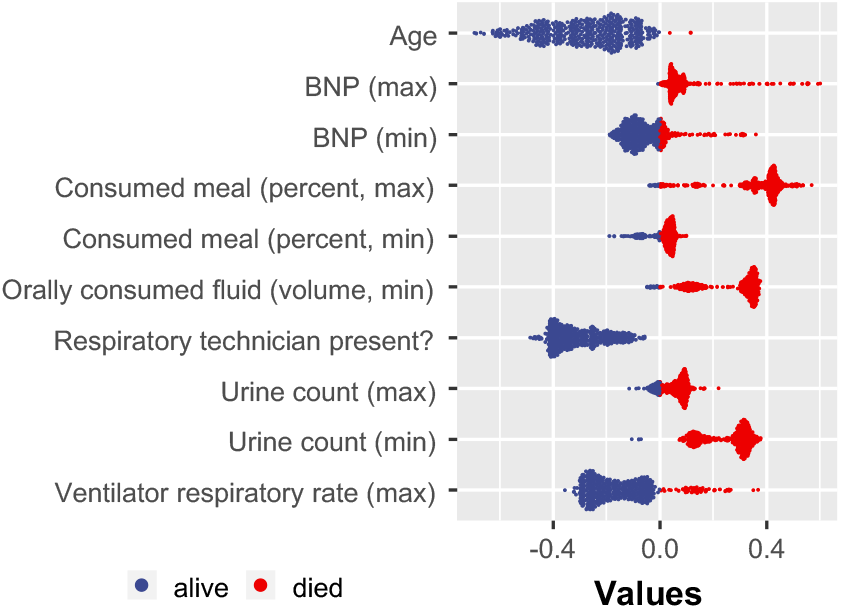
Example of SHAP value distribution for all of the tested patients. The visualization was created based on one of the trained XGBoost classifiers.

### 3.4 Discovered predictive features

Both feature-selection strategies identified a number of clinical characteristics that reflect overall patient health, cognitive status, and hospitalization risks associated with the patient’s condition. Here, we list some of the identified features, with a particular focus on those discovered by the LR-ENET model, which demonstrated superior AUC, sensitivity, and specificity compared to the XGBoost approach.

**CNS and cognition-related features**. A set of features related to the patient’s neurologic status was selected. The LR-ENET approach generated a model richer in these features than the XGBoost model.

- **Inability to assess the patient’s speech** may be caused by neurologic symptoms (such as loss of consciousness or the presence of delirium), but it may also be related to intubation. This EHR observation represents a surrogate measurement of the patient state in both instances, and its existence significantly suggests an elevated mortality risk.
- **Unclear and slurred speech** observed by nurses or physicians could be a key indicator of delirium [33]. Delirium is related to adverse outcomes during hospitalization (e.g., increased risk of complications) in post-acute care settings, and long-term follow-up (e.g., prolonged cognitive and functional impairment). The lack of speech clarity is only a surrogate measure for delirium that might be considered low value given that the presence of delirium is another explicitly defined feature in the analyzed dataset. However, it should be noted that there is a significant rate of under-recognition and lack of documentation of delirium in admitted patients, with less than 3% of cases documented by International Classification of Diseases-9 (ICD-9) codes in patients’ medical records [33, 34]
- **Any evidence of delirium**. Among the delirium-related characteristics included in the dataset is the expressly stated “Presence of delirium.” However, the feature selected as predictive was “Inability to assess delirium,” which may appear odd and counterintuitive. Examining the explicit delirium feature reveals a biserial correlation between the feature and the patient’s registered death of only 0.13, whereas the inability to assess delirium scores -0.25. This curious selection may be partially explained by the procedure required to assess delirium, which also explains the under-recognition of delirium in general. The Confusion Evaluation Method, the most used delirium assessment tool, requires an in-person, bedside discussion with the patient [35]. Because delirium fluctuates, interview-based approaches may overlook delirium that occurs beyond bedside interviews. It has been reported that manual searches of all records (e.g., nursing and physician notes, discharge summaries) may allow the determination of signs of delirium despite the lack of explicit notes in the records [33]
- **“Awake and able to response”** is another important neurologic assessment feature. A patient is scored positively if considered “awake,” “able to respond” (i.e., responding appropriately), and “oriented” (aware of self, place, and time) [36]. However, the patient’s situation may change on the first day after admission. Therefore, patients who were sedated and unable to be subjected to the neurologic examination may demonstrate full-strength cognitive abilities later. Thus, yet again, this feature provides value in combination with other features and cannot be considered in isolation. Interestingly, the Glasgow coma scale (GCS), which a measure used to determine the level of consciousness in trauma or critically ill individuals with impaired consciousness and which was also available in the dataset, has not been utilized by the models.

**Patient frailty-related features**. Several identified features described the patient’s overall condition and frailty upon hospital admission.

- **Braden score** describes the frailty of the patients. The Braden scale was created to identify early pressure, sore-prone patients. Six sub-scales of the score measure sensory perception, skin wetness, activity, mobility, friction and shear, and nutrition [37]. Although ample evidence exists for the usefulness and applicability of the Braden scale in predicting patients’ conditions during hospitalization [38], the scale has been criticized for lacking explainability and detail from the machine learning perspective [39]. On the other hand, a retrospective study of 146 COVID-19 patients demonstrated that the Braden score is indeed helpful for risk stratification at hospital admission, as the mortality among patients with BS *≤*15 was significantly higher than in patients with BS*>*15 [40]. In our research context, Braden score features also appear to identify particularly vulnerable patients. Interestingly, only two sub-scales (moisture and nutrition) have been included by the LR classifier in the final model. On the other hand, the XGBoost model did not rely on Braden scores.
- **Number of lumens** Multiple studies have demonstrated a substantial correlation between the number of PICC lumens and the risk of complications, including central-line associated bloodstream infection (CLABSI), venous thrombosis, and catheter occlusion [41, 42].
- **The requirement for a respiratory technician** to be present during the transportation of the patient is yet another predictive EHR feature that communicates the severity of the patient’s condition.
- **The urine voiding count feature** is connected with the lower urinary tract symptoms. The medical literature describes an association between LUTS and COVID [43–45]. According to these reports, there was a high prevalence of abnormal urinary storage symptoms, urine frequency, urgency, and urinary incontinence among the SARS-CoV-2-infected patients. The data indicate that the majority of COVID-19 patients may experience increased urination frequency, nocturia, and urgency during the infection. Also, patients with urine storage symptoms were found to have considerably higher COVID-19 severity levels than those without urine storage symptoms [45]. The urinary symptoms might be caused directly by inflammation or indirectly by COVID-19-related general dysfunction in the autonomic nervous system [46].

**Clinical laboratory test results** report on the patient’s physiological status. Specifically, two test results were identified by our feature selection methods.

- **The measurement of urea nitrogen in serum or plasma** (BUN SerPl test) assesses the kidney’s function. High urea nitrogen levels in the BUN test indicate problems with renal function or a reduction in blood supply to the kidneys. A reduction in urea nitrogen, as measured by the BUN test, indicates serious liver illness or malnutrition. The outcomes of the BUN test were put into the XGBoost model’s identified collection of features. However, they were not among the top 20 features identified by LR-ENET. In the analyzed cohort, there was a significant difference in test findings between patients who died and those who survived (*p*<0.001). These observations are consistent with literature reports [47, 48].
- **Brain natriuretic peptide (BNP)** is an active fragment (1-32) of the cardiac cell-produced ProBNP. It is elevated in right-sided and left-sided heart failure, as well as systolic and diastolic heart failure. It is therefore used to identify and treat heart failure. BNP test was recognized as an important feature by the XGBoost-feature selection, but not the LR-ENET. In the tested cohort, the values of the BNP test were substantially higher for male patients who died (*p*<0.001) but not for females (*p*=0.23). Others have recently postulated that BNP should be considered an early predictor of clinical severity in patients with COVID-19 pneumonia [49].

## 4 Discussion

When analyzing disease mortality causes, the search for predictive factors typically begins with the formulation of a hypothesis based on domain knowledge of the underlying diseases and initial preliminary evidence, such as case studies and anecdotal reports. This hypothesis-driven process is philosophically well-established and operationally widely accepted. Despite the fact that this conventional path of hypothesis-driven research has been challenged numerous times in recent years, particularly by the rise of genomics, for many researchers it is virtually synonymous with the scientific method itself [50]. The alternative paradigm, often referred to as data-driven research, begins with an agnostic stance that does not involve a preconceived hypothesis and instead employs either a data reduction process that results in the emergence of a model or a supervised model-building process that reveals predictive features that explain observed outcomes.

Here, we examined three methods for identifying factors predictive of mortality among COVID-19 patients with severe disease necessitating hospitalization. As a baseline, we utilized a standard statistical technique for formulating a hypothesis and generating or selecting hypothesized predictive factors through a GLM framework. In carrying out this method, we used a composite delirium factor, which is a combination of multiple EHR characteristics/symptoms associated with delirium occurrence. We also accounted for age, race, and sex, which we recognized as notable confounding variables plausibly related to the outcome. The developed model revealed a significant increase in the likelihood of death for hospitalized patients exhibiting any symptom of delirium.

Subsequently, we developed two feature discovery models using two well-established machine learning techniques. The first model utilized regularized elastic-net logistic regression. A sparse collection of predictors was generated using the model’s inbuilt feature selection capability. However, as previously established, the model was unstable, and the selection of reliably predictive features necessitated many model runs and a compilation of the findings. The most consistently predicted variables across numerous runs were evaluated using the SHAP method to obtain insight into their local significance for patient classification.

The second machine-learning model involved the use of XGBoost tree learning. In contrast to the sparsity of the LR-ENET method, this approach freely utilized all of the available clinical features. Throughout numerous rounds of independently run training and cross-validation, the algorithm demonstrated significantly higher stability, ultimately producing very similar results. To represent the feature importance, data from the internals of trained classifiers (gain and frequency) were extracted. A secondary XGBoost model was trained using the top features, and its SHAP values were assessed.

Our findings demonstrate that the two distinct classifiers relied on very different sets of predictive features during optimization and training. Age, surrogate measures for the patients’ cognitive status (neurological observations), features broadly describing the patients’ overall condition upon admission (such as the Braden score), and features associated with a risk of serious complications requiring hospitalization (such as the number of catheter lumens) were the descriptors that the LR-ENET model selected. It is interesting to note that common clinical laboratory test results were not chosen during the feature selection process by LR-ENET. XGBoost, on the other hand, placed considerable emphasis on laboratory test results, including values obtained from the BUN and BNP tests. XGBoost captured the patient’s overall condition by analyzing variables such as oral fluid consumption, oxygen saturation, ventilator use, etc. Despite the fact that the features chosen by XGBoost are frequently more quantitative and objective, the overall performance of the XGBoost model was marginally inferior in terms of specificity (such as in the case of the laboratory test results).

## 5 Conclusions

Risk stratification of hospitalized COVID-19 patients is crucial for informing individual treatment decisions that also account for resource allocation. Multiple risk models proposed so far have been based on mechanistic hypotheses regarding the SARS-CoV-2 mode of action, the association between comorbidities and observed outcomes, as well as hypothesized models of disease progression. In this report, we demonstrated the use of a machine learning-based, hypothesis-agnostic methodology for the discovery of predictive risk factors, which produces an easily understandable, observable, explainable, and actionable set of clinical features that may cause or be closely associated with in-hospital COVID-19 mortality. Our findings suggest that analyses of these features should be prioritized to identify patients with COVID-19 (and, potentially, other forms of acute respiratory distress syndrome) having an elevated risk of mortality.

## Data Availability

Access to the unprocessed clinical data can be obtained through the Regenstrief Institute in Indianapolis, IN, USA, after fulfilling the required agreements.

## Ethics statement

The study in question, identified by IRB protocol 2004316653, was approved by the Indiana University Institutional Review Board. As the study procedures involved the use of a deidentified electronic medical record database, consent was not required.

## References

[1] Zhou, Y. et al. Comorbidities and the risk of severe or fatal outcomes associated with coronavirus disease 2019: A systematic review and meta-analysis. International Journal of Infectious Diseases 99, 47–56 (2020).

[2] Mesas, A. E. et al. Predictors of in-hospital COVID-19 mortality: A comprehensive systematic review and meta-analysis exploring differences by age, sex and health conditions. PLOS ONE 15, e0241742 (2020).

[3] Belsky, J. A. et al. COVID-19 in immunocompromised patients: A systematic review of cancer, hematopoietic cell and solid organ transplant patients. Journal of Infection 82, 329–338 (2021).

[4] Li, C., Liu, J., Lin, J. & Shang, H. COVID-19 and risk of neurodegenerative disorders: A Mendelian randomization study. Translational Psychiatry 12, 1–6 (2022).

[5] Frontera, J. A. et al. Comparison of serum neurodegenerative biomarkers among hospitalized COVID-19 patients versus non-COVID subjects with normal cognition, mild cognitive impairment, or Alzheimer’s dementia. Alzheimer’s & Dementia 18, 899–910 (2022).

[6] Taquet, M., Geddes, J. R., Husain, M., Luciano, S. & Harrison, P. J. 6-month neurological and psychiatric outcomes in 236 379 survivors of COVID-19: A retrospective cohort study using electronic health records. The Lancet Psychiatry 8, 416–427 (2021).

[7] Farhadian, S. F., Seilhean, D. & Spudich, S. Neuropathogenesis of acute coronavirus disease 2019. Current Opinion in Neurology 34, 417–422 (2021).

[8] Thepmankorn, P. et al. Cytokine storm induced by SARS-CoV-2 infection: The spectrum of its neurological manifestations. Cytokine 138, 155404 (2021).

[9] Smith, M. & Alvarez, F. Identifying mortality factors from machine learning using shapley values – a case of COVID19. Expert Systems with Applications 176, 114832 (2021).

[10] Booth, A. L., Abels, E. & McCaffrey, P. Development of a prognostic model for mortality in COVID-19 infection using machine learning. Modern Pathology 34, 522–531 (2021).

[11] Ramón, A. et al. eXtreme Gradient Boosting-based method to classify patients with COVID-19. Journal of Investigative Medicine 70, 1472–1480 (2022).

[12] Vaid, A. et al. Machine learning to predict mortality and critical events in a cohort of patients with COVID-19 in new york city: Model development and validation. Journal of Medical Internet Research 22, e24018 (2020).

[13] Bertsimas, D. et al. COVID-19 mortality risk assessment: An international multi-center study. PLOS ONE 15, e0243262 (2020).

[14] Cavallaro, M., Moiz, H., Keeling, M. J. & McCarthy, N. D. Contrasting factors associated with COVID-19-related ICU admission and death outcomes in hospitalised patients by means of Shapley values. PLOS Computational Biology 17, e1009121 (2021).

[15] Banerjee, T., Paul, A., Srikanth, V. & Strümke, I. Causal connections between socioeconomic disparities and COVID-19 in the USA. Scientific Reports 12, 15827 (2022).

[16] Ikram, M., Shaikh, N. F., Vishwanatha, J. K. & Sambamoorthi, U. Leading predictors of COVID-19-related poor mental health in adult Asian Indians: An application of extreme gradient boosting and Shapley additive explanations. International Journal of Environmental Research and Public Health 20, 775 (2023).

[17] Bottino, F. et al. COVID mortality prediction with machine learning methods: A systematic review and critical appraisal. Journal of Personalized Medicine 11, 893 (2021).

[18] Hoerl, A. E. & Kennard, R. W. Ridge regression: Biased estimation for nonorthogonal problems. Technometrics 12, 55–67 (1970).

[19] Tibshirani, R. Regression shrinkage and selection via the LASSO. Journal of the Royal Statistical Society: Series B (Methodological) 58, 267–288 (1996).

[20] Zou, H. & Hastie, T. Regularization and variable selection via the elastic net. Journal of the Royal Statistical Society: Series B (Statistical Methodology) 67, 301–320 (2005).

[21] Jenul, A. et al. RENT—repeated elastic net technique for feature selection. IEEE Access 9, 152333–152346 (2021). 2009.12780.

[22] Nogueira, S., Sechidis, K. & Brown, G. On the stability of feature selection algorithms. Journal of Machine Learning Research 18, 1–54 (2018).

[23] Xu, H., Caramanis, C. & Mannor, S. Sparse algorithms are not stable: A no-free-lunch theorem. IEEE Transactions on Pattern Analysis and Machine Intelligence 34, 187–193 (2012).

[24] Menardi, G. & Torelli, N. Training and assessing classification rules with imbalanced data. Data Mining and Knowledge Discovery 28, 92–122 (2014).

[25] Chen, T. & Guestrin, C. XGBoost: A scalable tree boosting system. In Proceedings of the 22nd ACM SIGKDD International Conference on Knowledge Discovery and Data Mining, KDD ‘16, 785–794 (ACM, New York, NY, USA, 2016). URL http://doi.acm.org/10.1145/2939672.2939785.

[26] Shapley, L. S. A value for n-person games. In Roth, A. E. (ed.) The Shapley Value: Essays in Honor of Lloyd S. Shapley, 31–40 (Cambridge University Press, Cambridge, 1988).

[27] Song, E., Nelson, B. L. & Staum, J. Shapley effects for global sensitivity analysis: Theory and computation. SIAM/ASA Journal on Uncertainty Quantification 4, 1060–1083 (2016).

[28] Owen, A. B. & Prieur, C. On Shapley value for measuring importance of dependent inputs. SIAM/ASA Journal on Uncertainty Quantification 5, 986–1002 (2017).

[29] Lundberg, S. M. & Lee, S.-I. A unified approach to interpreting model predictions. Advances in neural information processing systems 30 (2017).

[30] Aas, K., Jullum, M. & Løland, A. Explaining individual predictions when features are dependent: More accurate approximations to Shapley values. Artificial Intelligence 298, 103502 (2021).

[31] Pearl, J. Causality: Models, Reasoning and Inference (Cambridge University Press, Cambridge, U.K.; New York, 2009), 2nd edition edn.

[32] Janzing, D., Minorics, L. & Bloebaum, P. Feature relevance quantification in explainable AI: A causal problem. In Proceedings of the Twenty Third International Conference on Artificial Intelligence and Statistics, 2907–2916 (PMLR, 2020).

[33] Puelle, M. R. et al. The language of delirium: Keywords for identifying delirium from medical records. Journal of Gerontological Nursing 41, 34–42 (2015).

[34] Inouye, S. K. et al. A chart-based method for identification of delirium: Validation compared with interviewer ratings using the confusion assessment method. Journal of the American Geriatrics Society 53, 312–318 (2005).

[35] Inouye, S. K. et al. Clarifying confusion: The confusion assessment method. Annals of Internal Medicine 113, 941–948 (1990).

[36] Shahrokhi, M. & Asuncion, R. M. D. Neurologic Exam. In StatPearls (StatPearls Publishing, Treasure Island (FL), 2022).

[37] Bergstrom, N., Braden, B. J., Laguzza, A. & Holman, V. The Braden scale for predicting pressure sore risk. Nursing Research 36, 205–210 (1987 Jul-Aug).

[38] Brown, S. J. The Braden Scale: A review of the research evidence. Orthopaedic Nursing 23, 30–38 (2004).

[39] Alderden, J. et al. Explainable artificial intelligence for predicting hospital-acquired pressure injuries in COVID-19–positive critical care patients. Computers, Informatics, Nursing 40, 659–665 (2022).

[40] Lovicu, E., Faraone, A. & Fortini, A. Admission Braden scale score as an early independent predictor of in-hospital mortality among inpatients with COVID-19: A retrospective cohort study. Worldviews on Evidence-Based Nursing 18, 247–253 (2021).

[41] Chopra, V. et al. Risk of venous thromboembolism associated with peripherally inserted central catheters: A systematic review and meta-analysis. The Lancet 382, 311–325 (2013).

[42] Bozaan, D. et al. Less lumens-less risk: A pilot intervention to increase the use of single-lumen peripherally inserted central catheters. Journal of Hospital Medicine 14, 42–46 (2019).

[43] Mumm, J.-N. et al. Urinary frequency as a possibly overlooked symptom in COVID-19 patients: Does SARS-CoV-2 cause viral cystitis? European Urology 78, 624–628 (2020).

[44] Swatesutipun, V. & Tangpaitoon, T. Lower urinary tract symptoms (LUTS) related to COVID-19: Review article. Journal of the Medical Association of Thailand 104, 1045–9 (2021).

[45] Swatesutipun, V. & Tangpaitoon, T. The prevalence and risk factors of storage urinary symptoms in symptomatic COVID-19 patients who were treated in cohort ward and field hospital. Siriraj Medical Journal 74, 134–141 (2022).

[46] Buoite Stella, A. et al. Autonomic dysfunction in post-COVID patients with and witfhout neurological symptoms: A prospective multidomain observational study. Journal of Neurology 269, 587–596 (2022).

[47] Ok, F., Erdogan, O., Durmus, E., Carkci, S. & Canik, A. Predictive values of blood urea nitrogen/creatinine ratio and other routine blood parameters on disease severity and survival of COVID-19 patients. Journal of Medical Virology 93, 786–793 (2021).

[48] Cheng, A. et al. Diagnostic performance of initial blood urea nitrogen combined with D-dimer levels for predicting in-hospital mortality in COVID-19 patients. International Journal of Antimicrobial Agents 56, 106110 (2020).

[49] Cilingir, B. M., Askar, S., Meral, A. & Askar, M. Can B-type natriuretic peptide (BNP) levels serve as an early predictor of clinical severity in patients with COVID-19 pneumonia. Clinical laboratory 68 (2022).

[50] Defining the scientific method. Nature Methods 6, 237–237 (2009).

